# Common Acute Pesticide Poisoning in a Single Centre Study in Bangladesh

**DOI:** 10.1101/2025.03.24.25324472

**Authors:** Sabbiha Nadia Majumder, Md Robed Amin, Md Jahid Hasan, Mohammad Rafiqul Islam, M A Faiz

**Author notes:** Corresponding Authors: Sabbiha Nadia Majumder; Md Robed Amin. All authors contributed to the study conception, design, data collection, and manuscript preparation. All authors read and approved of the final manuscript. The datasets used and/or analyzed during the current study are available from the corresponding author upon reasonable request. Ethical approval for this study was obtained from the Institutional Review Board of Dhaka Medical College Hospital.

## Abstract

**Background:** Acute pesticide poisoning is a primary public health concern in Bangladesh, contributing to significant morbidity and mortality, primarily due to deliberate self-harm. Accurate and timely identification of the ingested pesticide is essential for effective management, but this remains challenging due to limited toxicological resources and inadequate information at the point of care.

**Methods:** A cross-sectional observational study was conducted at Dhaka Medical College Hospital from June 2017 to February 2018. Patients aged 18 years and older presenting with features of acute pesticide poisoning were enrolled if they could provide a sample of the ingested substance, either physically or digitally (via photos or messaging apps). Clinical characteristics, toxidromes, and outcomes were recorded using a standardized case record form. Data was analyzed using SPSS version 17 to identify toxicological patterns and predictors of mortality.

**Results:** Among 236 patients, organophosphate compounds (OPCs) accounted for 52.2% of cases, followed by miticides (21.1%), herbicides (9.1%), and carbamates (3.4%). Intentional ingestion accounted for 93% of cases. The overall mortality rate was 12.7% (30 deaths), with OPCs responsible for 66.7% of fatalities. A Glasgow Coma Scale score below 9 (OR 12.12; p<0.001) and constricted pupils at admission were significant predictors of poor outcomes. Illiteracy was also associated with increased mortality (OR 3.78; p=0.002). Identifying the ingested agent through physical or digital samples substantially aided clinical decision-making.

**Conclusion:** Acute pesticide poisoning in Bangladesh is primarily due to OPCs and affects predominantly young adults. Collection of ingested agent samples, including digital images, can facilitate targeted treatment and improve outcomes in resource-limited healthcare settings.

## Introduction

Acute pesticide poisoning represents a significant public health concern in Bangladesh, accounting for a substantial proportion of emergency visits and hospital admissions. It is the seventh leading cause of hospital mortality nationwide^1^. Pesticide ingestion remains the most common method of intentional poisoning across many Asian countries ^2^. Regional studies from southern and hilly areas of Bangladesh have consistently identified pesticides as the predominant agents in poisoning incidents ^3,4,5^.

The increasing availability and accessibility of agrochemicals have made pesticide poisoning a preventable yet persistent cause of illness and death. Pesticides are divided into organophosphates (OPs) and non-organophosphates (non-OPs), each with unique toxicological profiles and treatment approaches. However, clinical management is often complicated by a lack of information about the specific substance ingested, limited access to toxicological analysis, and insufficient awareness among healthcare providers and the community.

This hospital-based cross-sectional study was conducted to identify the specific types of pesticides ingested by patients presenting with acute poisoning and to evaluate their short-term clinical outcomes. The study aims to provide insight into the current situation of pesticide poisoning in a tertiary care hospital in Bangladesh and underscore the urgent need for improved diagnostic practices, community awareness, and treatment protocols.

## Study Objectives

### Primary Objective

- To identify the pesticide agents involved in acute poisoning cases through collected physical or digital specimens (e.g., bottles, containers, or images) presented by patients or their attendants at a tertiary care hospital in Bangladesh.

### Secondary Objectives

- To classify the types of pesticides involved in poisoning cases, including insecticides, herbicides, acaricides, fungicides, and rodenticides.
- To evaluate the correlation between clinical toxidromes and the categories of pesticides ingested.

## Materials and Methods

### Study Site and Setting

This study was conducted in all adult medicine units of Dhaka Medical College Hospital (DMCH), a tertiary care facility located in Dhaka, Bangladesh. Data were collected from 236 patients diagnosed with acute pesticide poisoning during the study period.

### Study Design and Duration

This was a hospital-based, cross-sectional observational study that lasted nine months, from July 1, 2017, to March 31, 2018.

### Study Population

All adult patients aged 18 years and older who presented with clinical features suggestive of acute pesticide poisoning were eligible for inclusion. Patients with either a confirmed history of pesticide ingestion or those displaying supportive clinical toxidromes were considered for the study. Individuals under 18 years of age were excluded.

### Pesticide Identification

Pesticides involved in each case were identified using a reference list of registered agricultural and public health pesticides in Bangladesh, published by the Bangladesh Crop Protection Association in 2008. This list included information on active ingredients, brand names, trade names, manufacturers, and registration details.

### Data Collection Procedure

After obtaining informed written consent, clinical data was collected using a pretested case record form (CRF). A trained research assistant (RA) conducted the initial clinical evaluation, which was subsequently verified by one of the investigators. Patients were included regardless of the intent behind ingestion (deliberate self-harm or accidental). Clinical findings were documented, including presenting symptoms, vital signs, level of consciousness, and toxidromes.

Patients or their attendants were asked to provide a physical specimen (e.g., a container or label) or a digital image of the ingested pesticide via mobile applications such as Viber, Facebook Messenger, IMO, or email. Only cases where a sample or image was successfully submitted were included in the final analysis.

Each specimen or image was reviewed for labeling details, including the active ingredient, manufacturer, trade name, and branding. The RA and a co-investigator cross-verified the product using the official pesticide registration list maintained by the Plant Protection Wing, Department of Agricultural Extension, Khamarbari, Dhaka. In cases of uncertainty, the same agency reconfirmed the product’s identity.

Pesticides were further classified into categories: insecticides, herbicides, acaricides, fungicides, and rodenticides. Organophosphate (OP) compounds and carbamates were classified as agent types 1 and 2, respectively.

All physical specimens were securely stored in a locked locker in a designated room and handled with gloves within the Department of Medicine at DMCH for six months. They were then handed over to the Syngenta pharmaceutical team for disposal according to their protocol.

### Subject Selection

#### Inclusion Criteria

Patients were eligible for inclusion if they met the following conditions:

- Adults aged 18 years and above admitted to the Medicine units of Dhaka Medical College Hospital (DMCH) with clinical features suggestive of acute pesticide poisoning. Diagnosis was based on the presence of a clinical toxidrome particularly cholinergic features for suspected organophosphate poisoning and/or the identification of the ingested agent through a physical specimen (e.g., bottle, container) brought by the patient or their attendant.
- Patients with a confirmed history of acute pesticide ingestion, even in the absence of clinical symptoms, were included if the ingested substance could be identified through photographic or digital evidence (e.g., images sent via mobile applications or email), or by inspection of the physical product.

#### Exclusion Criteria

- Cases in which the submitted sample or image confirmed that the ingested agent was not a pesticide (e.g., pharmaceutical or household substance) were excluded.
- Patients or legal guardians who declined to provide written informed consent were not included in the study.

## Ethical Considerations, Data Collection, and Analysis

### Confidentiality and Data Protection

All collected data were securely stored in a locked cabinet within the study premises. Access to the data and source documents was restricted to the principal investigator (PI), co-investigators, and the designated research assistant (RA). Identifiable patient information was anonymized during data entry, and confidentiality was maintained throughout the study in compliance with ethical research standards. Data was disclosed only with the explicit permission of the participants.

### Informed Consent

Written informed consent was obtained from all study participants. Prior to enrollment, participants were informed of the study’s objectives, procedures, potential risks, and benefits. Consent was voluntary, and participants retained the right to withdraw at any point without prejudice. In cases where the patient was unable to provide consent due to clinical condition, consent was obtained from a legally authorized representative (e.g., family member or guardian). No individuals under 18 years of age were included in the study, in accordance with the exclusion criteria.

### Data Collection and Quality Control

Data were collected using a standardized and pre-tested case record form (CRF). The research assistant was formally trained in data collection procedures. Each patient’s clinical profile was recorded at admission and verified by the PI or a co-investigator. To ensure consistency and accuracy, a one-day training workshop was held for the study team. The PI regularly monitored the data collection process and performed random checks to ensure data integrity.

### Data Management and Statistical Analysis

Data were entered and cleaned using SPSS software (version 16, SPSS Inc., Chicago, IL, USA). Descriptive statistics were used to summarize patient demographics and clinical characteristics. The Kruskal–Wallis and Wilcoxon rank sum tests were applied to assess associations between categorical and ordinal variables. Comparison between organophosphate and non-organophosphate poisoning cases was conducted using paired t-tests. The p-value of <0.005 was considered statistically significant. Mortality outcomes were evaluated using odds ratios with corresponding 95% confidence intervals.

## Results

In the study, 236 patients were enrolled, and organophosphate (OP) compounds were identified as the most frequently implicated agents, accounting for 52.2% of cases. Non-OP pesticides included miticides (21.1%), herbicides (9.1%), carbamates (3.4%), and rodenticides (2.6%). The age group most affected was 15 to 25 years, although the highest fatality rate was observed among patients older than 40 years.

A total of 30 deaths (12.7%) were recorded, with 20 of these (66.7%) attributed to the ingestion of OP compounds. Most poisoning incidents (93%) resulted from deliberate self-harm, while the remaining cases were either accidental or, in rare instances, homicidal. Factors such as a Glasgow Coma Scale (GCS) score of less than 9 upon admission and pupillary constriction were significantly associated with increased mortality. In contrast, initial vital signs, including heart rate and blood pressure, did not significantly correlate with patient outcomes.

Figure 1 presents representative images of pesticide containers and labels brought in by patients or their attendants, highlighting the utility of sample-based identification in clinical assessments.

**Fig 1:**
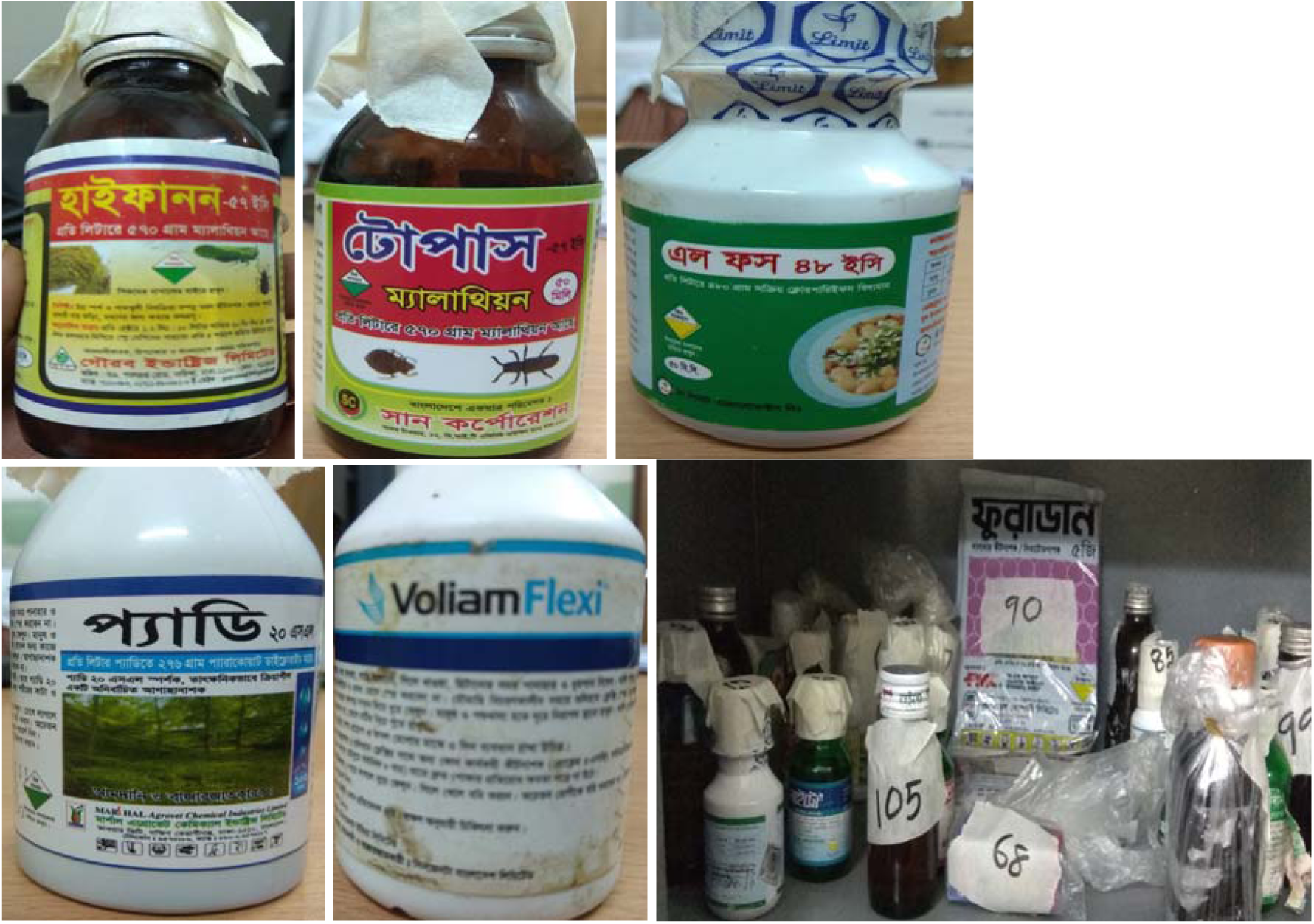
Brought samples of pesticide and their stored status in locker.

**Fig 2:**
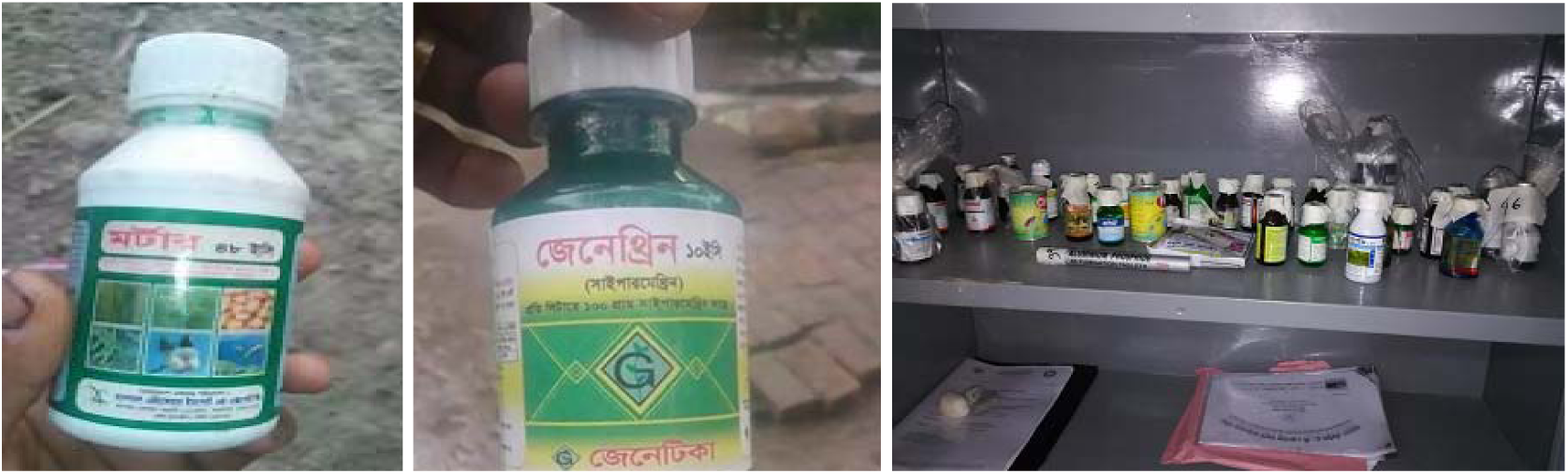
OP compounds that were brought to hospital premise by patients’ attendant and storage.

**Fig 3:**
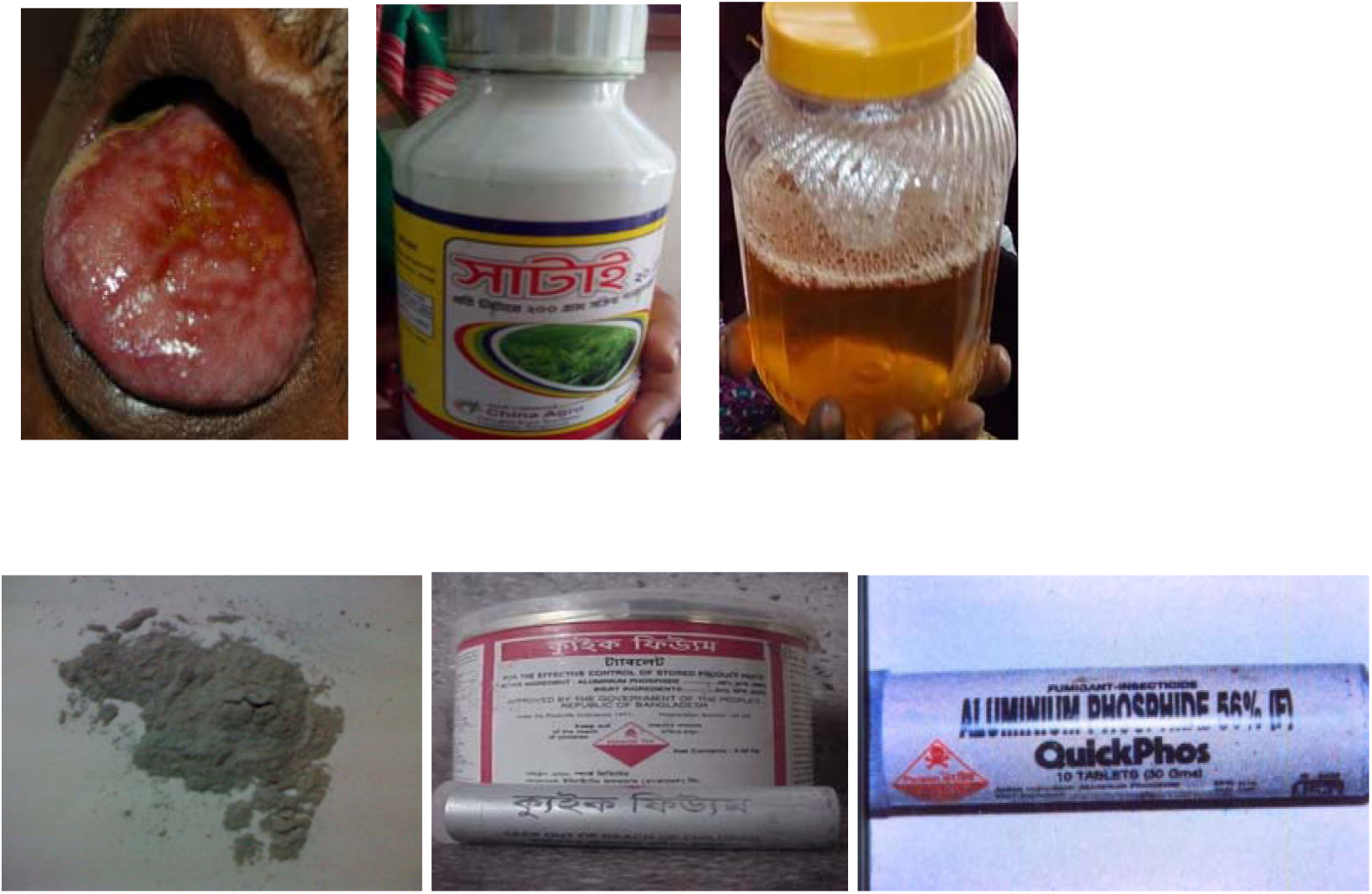
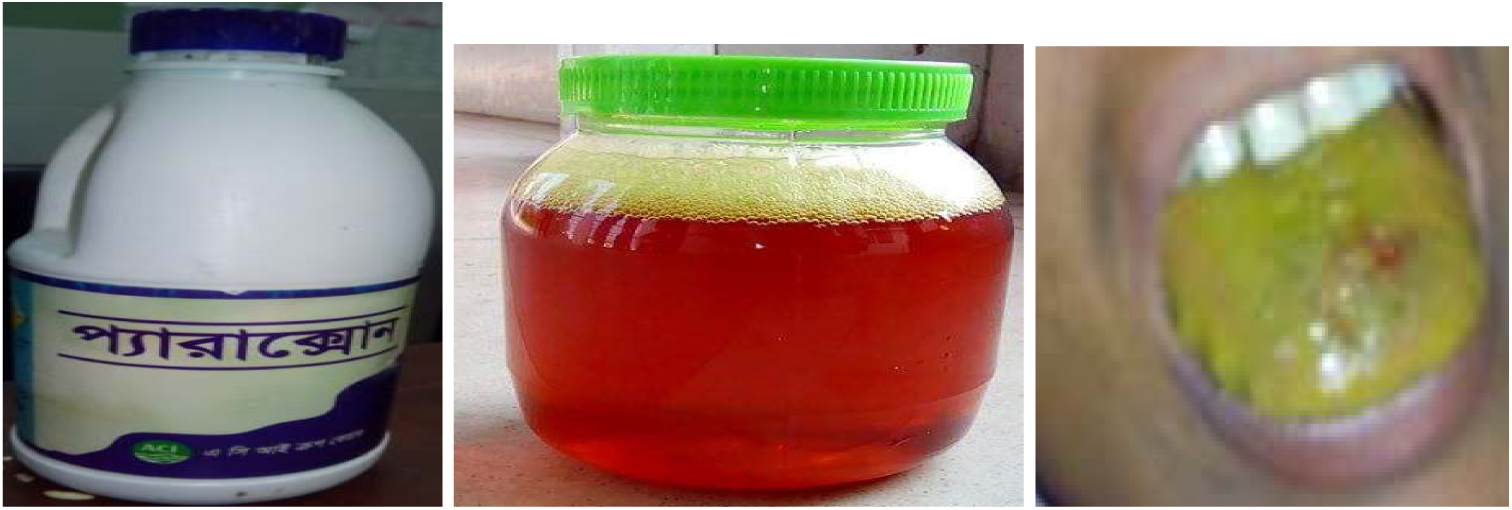
Samples of non-OP compound (Paraquat and Aluminum phosphide poisoning cases with patient’s clinical problems)

Further classification of the pesticides revealed that 52% of cases involved organophosphate (OP) compounds, while the remaining 48% were attributed to non-organophosphate (non-OP) agents (Pie chart 1). Among the non-OP group, the most common category was miticides or other pesticides, comprising 44% of non-OP cases. Herbicides accounted for approximately 21%, followed by stored grain pest control agents (15%), carbamates (8%), and fungicides (7%) (Pie Chart 2).

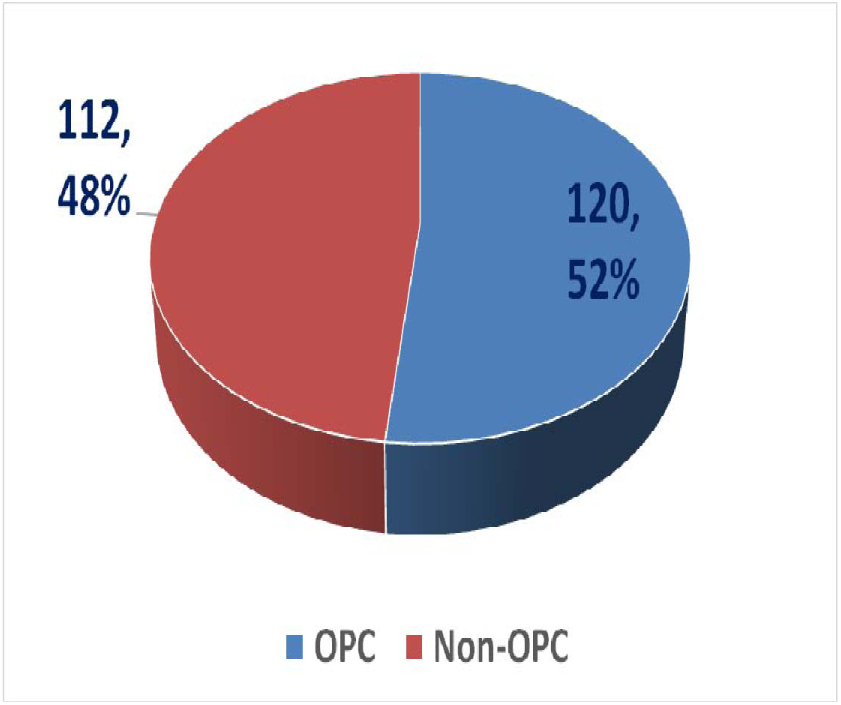
Pie chart 1: OP vs non-OP frequency.

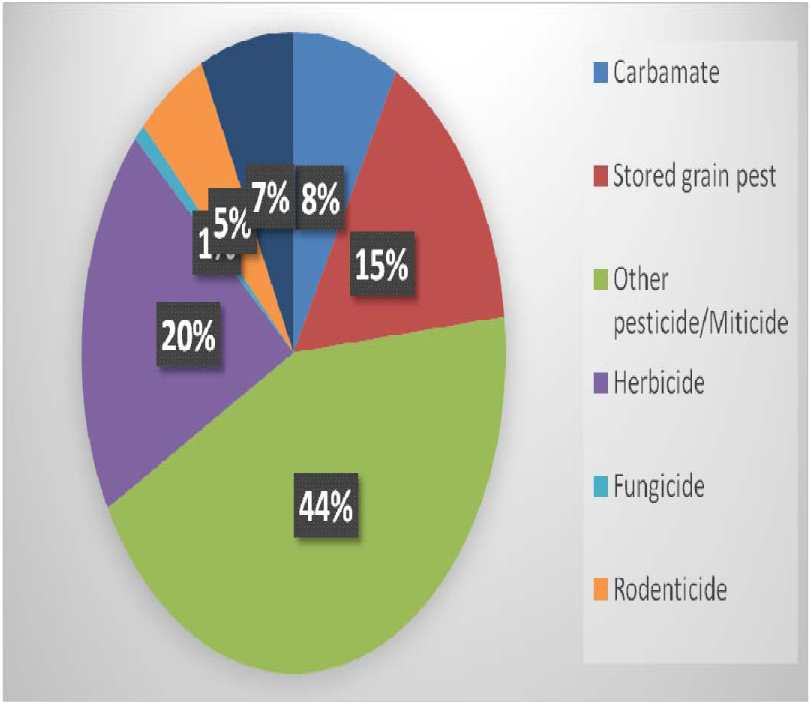
Pie Chart 2: Different classified Pesticide.

The relationship between patient age, type of pesticide exposure, and mortality was analyzed across the study population (Table 1 and line graph 1). The highest incidence of pesticide poisoning occurred in the 15–25-year age group, reflecting a trend of deliberate self-harm among younger individuals. Although most cases occurred in this demographic, the highest case fatality rate was observed in patients aged over 35 years. A secondary peak in the mortality rate was also noted among elderly individuals aged over 65 years.

**Table 1:**
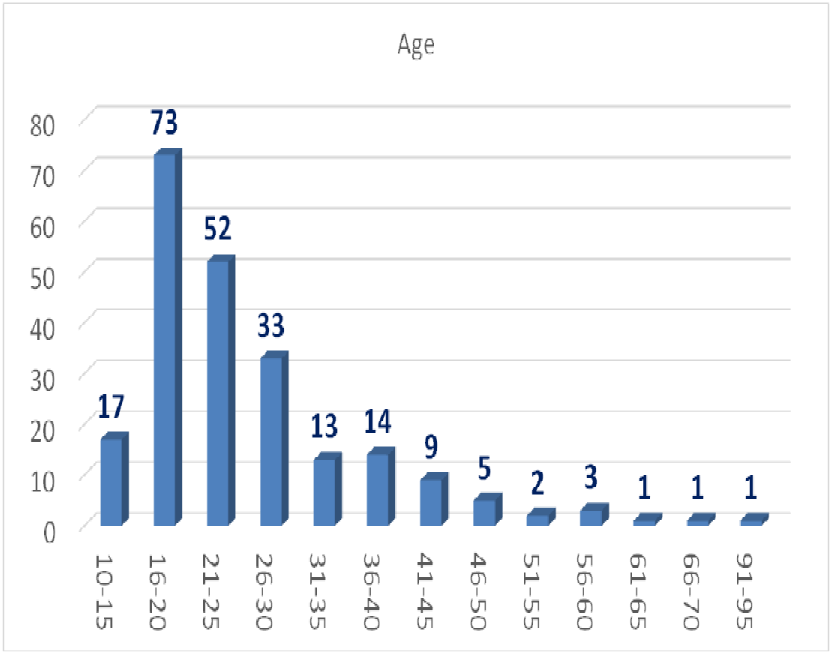
Age distribution of all cases.

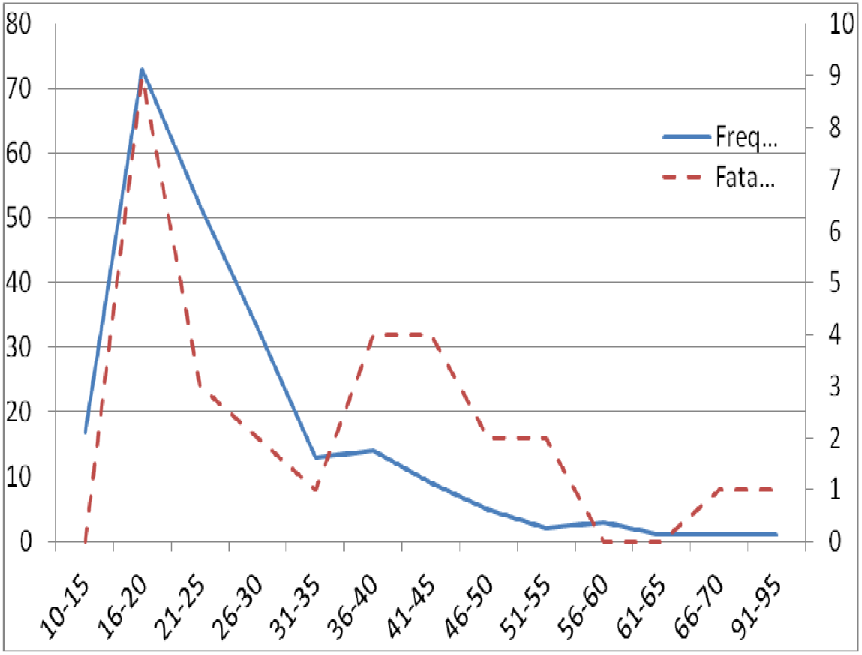
Line graph 1: OP vs non-OP outcome in relation to age

Among patients who attempted self-harm, most were rescued by family members or bystanders and subsequently taken to a healthcare facility. The time between ingestion and arrival at the hospital varied significantly. Notably, 86% of patients presented to medical facilities within 2 to 6 hours of the poisoning event (see Table 2). Timely intervention during this time frame may have contributed to reducing morbidity and mortality. In terms of initial care-seeking behavior, most patients (97.5%) were taken directly to government hospitals. Only a small percentage sought assistance from other providers, including private clinics, community-based practitioners, and traditional healers (refer to Table 3). This underscores the crucial role of public sector hospitals in managing acute poisoning cases in Bangladesh, likely due to factors such as accessibility, cost, and medicolegal considerations.

**Table 2:**
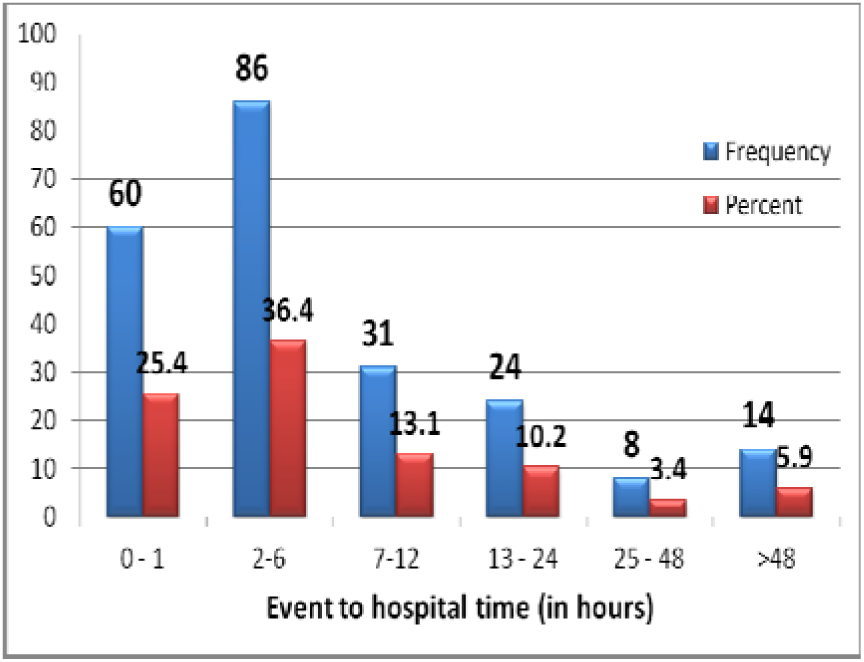
Event to hospital time in hours.

**Table 3:**
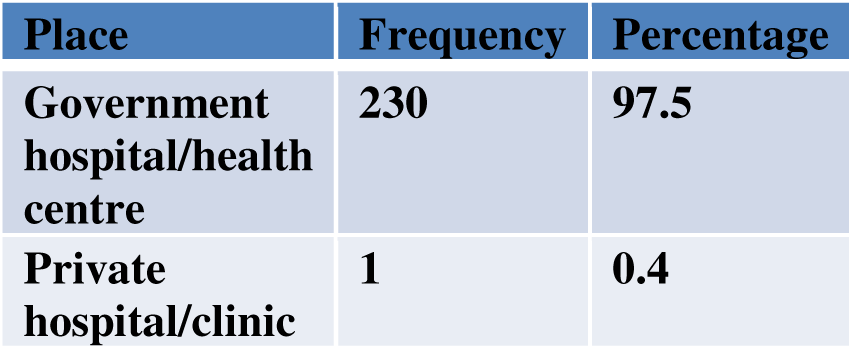
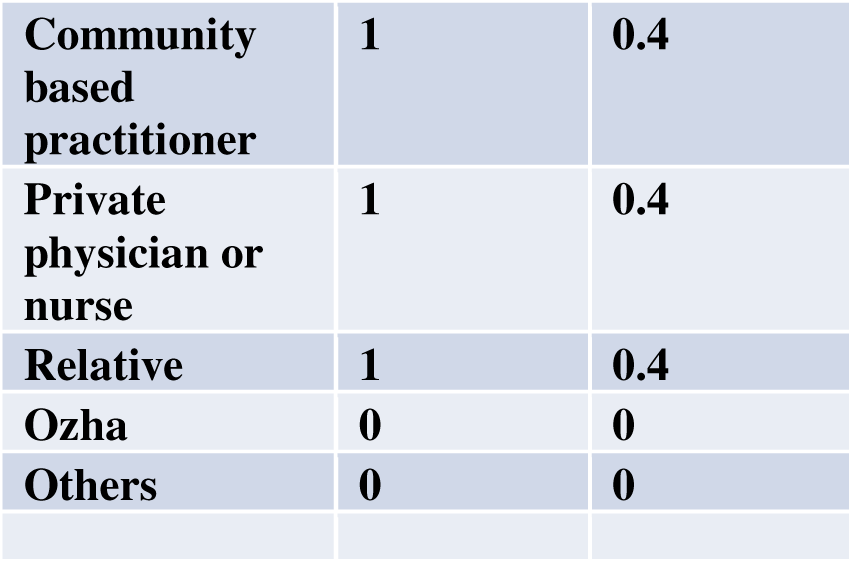
Where the patient was taken first.

| Place | Frequency | Percentage |
| --- | --- | --- |
| Government hospital/health centre | 230 | 97.5 |
| Private hospital/clinic | 1 | 0.4 |
| Community based practitioner | 1 | 0.4 |
| Private physician or nurse | 1 | 0.4 |
| Relative | 1 | 0.4 |
| Ozha | 0 | 0 |
| Others | 0 | 0 |

The clinical manifestations of pesticide poisoning in this study varied considerably depending on the type of agent involved. A broad spectrum of symptoms of differing intensity was documented across cases (Table 4). The most frequently reported symptoms included vomiting (76.7%), sweating (19.1%), respiratory distress (18.6%), and a burning sensation in the throat (15.7%). Other less common symptoms included salivation, abdominal pain, dizziness, myoclonus, and decreased consciousness. This heterogeneity in clinical presentation underscores the importance of early recognition of toxidromes and the need for substance-specific management strategies.

**Table 4:** Symptoms and signs of patients.

| Symptom | Frequency | Percentage |
| --- | --- | --- |
| Burning throat | 37 | 15.7 |
| Salivation | 16 | 6.8 |
| Nausea | 13 | 5.5 |
| Vomiting | 181 | 76.7 |
| Dysphagia | 3 | 1.3 |
| Abdominal Pain | 15 | 6.4 |
| Diarrhea | 13 | 5.5 |
| Sweating | 45 | 19.1 |
| Vertigo | 5 | 2.1 |
| Headache | 4 | 1.7 |
| Respiratory distress | 44 | 18.6 |
| Weakness of limb | 8 | 3.4 |
| Weakness of eye | 2 | 8 |
| Myoclonus | 2 | 8 |
| Fever | 3 | 1.3 |
| Poor consciousness | 4 | 1.7 |
| **Blood mixed saliva** | **1** | **0.4** |
| **Burning Chest** | **1** | **0.4** |
| **Difficulty in opening** | **1** | **0.4** |
| **Difficulty raising hands** | **1** | **0.4** |
| **Disorientation** | **2** | **0.8** |
| **Frothing** | **24** | **10.1** |
| **Generalized weakness** | **2** | **0.8** |
| **Haematemesis** | **2** | **0.8** |
| **Hiccup** | **1** | **0.4** |
| **Decreased urine** | **3** | **1.3** |
| **Hemoptysis** | **8** | **3.4** |
| **Jaundice** | **2** | **0.8** |
| **Loose motion** | **4** | **1.7** |
| **Muscle fasciculation** | **1** | **0.4** |
| **Restlessness** | **7** | **2.9** |
| **Convulsion** | **1** | **0.4** |

Several clinical parameters were found to be associated with increased mortality in cases of pesticide poisoning, particularly among patients exposed to organophosphate (OP) compounds. A Glasgow Coma Scale (GCS) score below 9 on admission was significantly correlated with fatal outcomes and emerged as a strong predictor of mortality (Table 5). Additionally, systolic blood pressure below 80 mmHg was more frequently observed among non-survivors in the OP group, further highlighting the prognostic relevance of hemodynamic status. These findings underscore the importance of early assessment of vital signs and neurological status in predicting clinical outcomes and guiding the urgency of intervention in acute pesticide poisoning.

**Table 5:** Factors predicting mortality in pesticide poisoning.

| **Variable** | **Total/**<br>**Missing**<br>**data** | **Category** | **Proport**<br>**ion of**<br>**deaths**<br>**(%)** | **Of All** |  | **Survivors** |  | **Deaths** |  | **Odds**<br>**ratio** | **95%**<br>**CI** | **P value** |
| --- | --- | --- | --- | --- | --- | --- | --- | --- | --- | --- | --- | --- |
|  |  |  | Among<br>all | N | % | N | % | N | % |  |  |  |
|  | Total<br>data<br>236 |  |  |  |  |  |  | Total<br>death<br>30 | 12.9 |  |  |  |
| Glasgow<br>Coma<br>Score | 232/4 | <9 | 33.33% | 18 | 7.8<br>% | 8 | 44.4<br>% | 10 | 55.6<br>% | 12.12 | 4.29 –<br>34.21 | <0.001 |
| Blood pressure | 232/4 | Systolic <80 | 16.7% | 16 | 6.9 % | 11 | 68.8 % | 5 | 31.2 % | 3.47 | 1.11 – 10.81 | 0.03 |
|  |  | Diastolic <60 | 3.0% | 33 | 14.2 % | 26 | 78.8 % | 7 | 21.2 % | 2.06 | 0.80 – 5.27 | 0.13 |
| Heart rate | 230/6 | >100 | 1.3% | 28 | 12.2 % | 25 | 89.3 % | 3 | 10.7 % | 0.81 | 0.22- 2.88 | 0.74 |
|  |  | <60 | 0.4% | 6 | 2.6 % | 5 | 83.3 % | 1 | 16.7 % | 1.4 | 0.15- 12.42 | 0.76 |
| Pupil | 223/13 | Normal | 4.9% | 11 8 | 52.9 % | 10 7 | 90.7 % | 11 | 9.3% | 1 (base) |  |  |
|  |  | Dilated | 1.8% | 22 | 9.9 % | 18 | 81.8 % | 4 | 18.2 % | 2.16 | 0.62 – 7.53 | 0.22 |
|  |  | Constricted | 6.7% | 83 | 37.2 % | 68 | 81.9 % | 15 | 18.1 % | 2.14 | 0.93 – 4.94 | 0.07 |
|  |  | Normal |  |  |  |  | Odds relative to other categories within the same variable |  |  | 0.46 | 0.21 – 1.03 | 0.05 |
|  |  | Dilated |  |  |  |  |  |  |  | 1.49 | 0.46 – 4.76 | 0.49 |
|  |  | Constricted |  |  |  |  |  |  |  | 1.83 | 0.84 – 3.98 | 0.12 |

Sociodemographic characteristics were also analyzed in relation to patient outcomes (Table 6). Illiteracy was identified as a significant predictor of mortality, with illiterate patients demonstrating higher odds of death compared to those with formal education (odds ratio [OR] 3.78; 95% confidence interval [CI]: 1.62–8.81; p = 0.002).

**Table 6:**
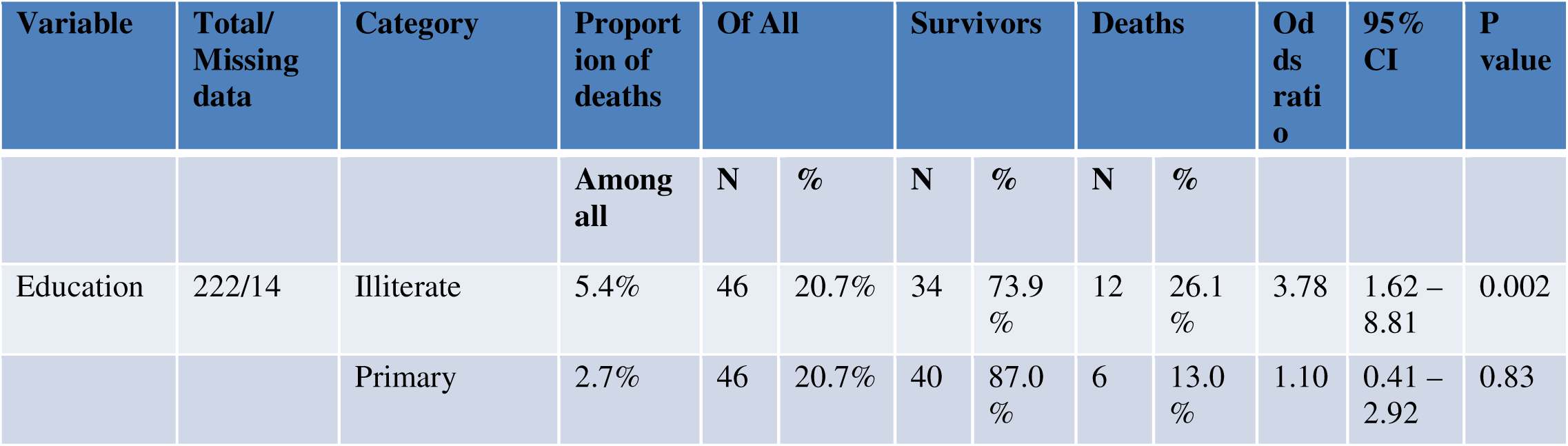
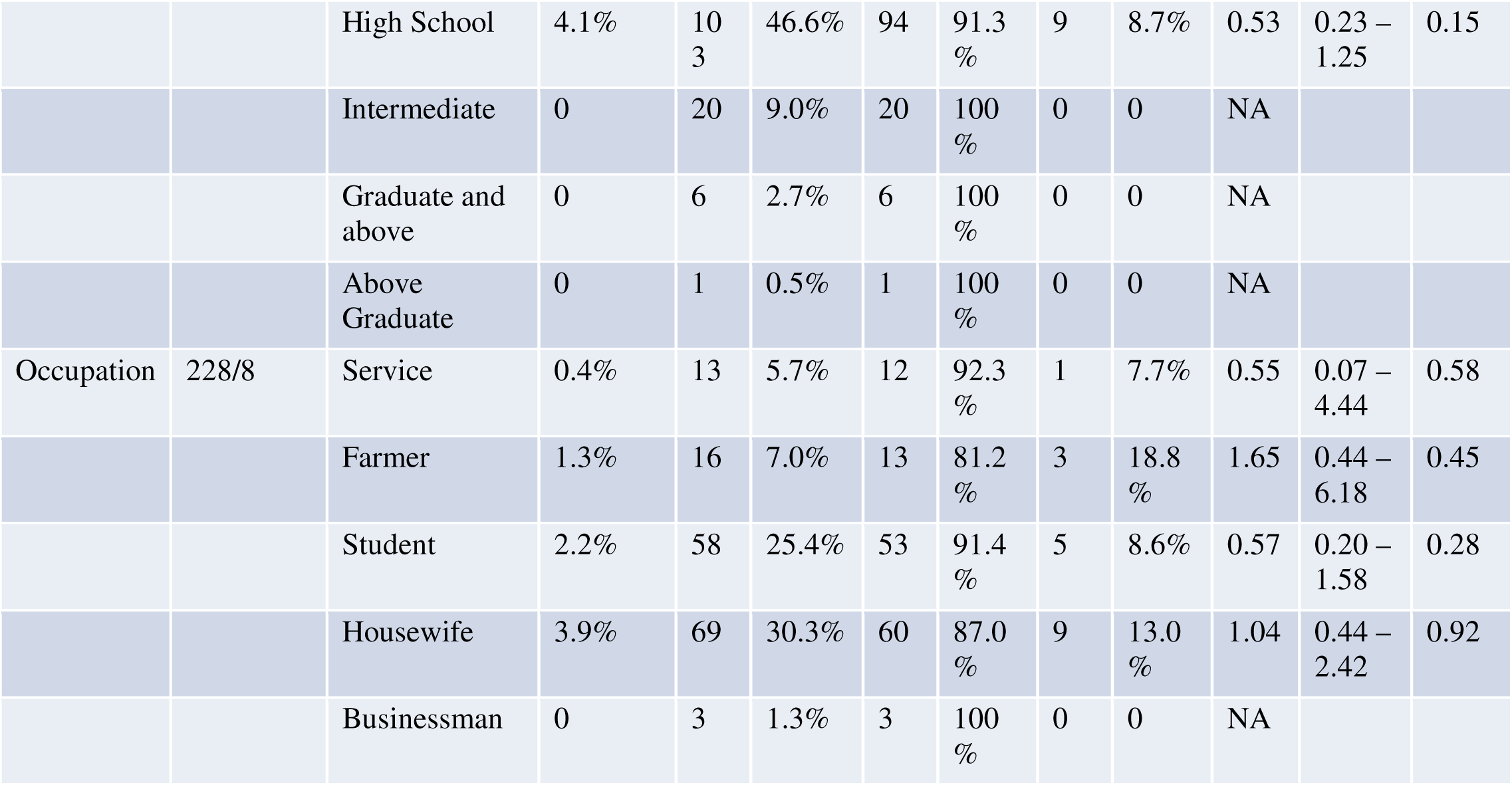
Education and occupation in relation to death of patients (n-30/236)

| Variable | Total/<br>Missing<br>data | Category | Proport<br>ion of<br>deaths | Of All |  | Survivors |  | Deaths |  | Od<br>ds<br>rati<br>o | 95%<br>CI | P<br>value |
| --- | --- | --- | --- | --- | --- | --- | --- | --- | --- | --- | --- | --- |
|  |  |  | Among<br>all | N | % | N | % | N | % |  |  |  |
| Education | 222/14 | Illiterate | 5.4% | 46 | 20.7% | 34 | 73.9<br>% | 12 | 26.1<br>% | 3.78 | 1.62 – 8.81 | 0.002 |
|  |  | Primary | 2.7% | 46 | 20.7% | 40 | 87.0<br>% | 6 | 13.0<br>% | 1.10 | 0.41 – 2.92 | 0.83 |
|  |  | High School | 4.1% | 10<br>3 | 46.6% | 94 | 91.3<br>% | 9 | 8.7% | 0.53 | 0.23 –<br>1.25 | 0.15 |
|  |  | Intermediate | 0 | 20 | 9.0% | 20 | 100<br>% | 0 | 0 | NA |  |  |
|  |  | Graduate and<br>above | 0 | 6 | 2.7% | 6 | 100<br>% | 0 | 0 | NA |  |  |
|  |  | Above<br>Graduate | 0 | 1 | 0.5% | 1 | 100<br>% | 0 | 0 | NA |  |  |
| Occupation | 228/8 | Service | 0.4% | 13 | 5.7% | 12 | 92.3<br>% | 1 | 7.7% | 0.55 | 0.07 –<br>4.44 | 0.58 |
|  |  | Farmer | 1.3% | 16 | 7.0% | 13 | 81.2<br>% | 3 | 18.8<br>% | 1.65 | 0.44 –<br>6.18 | 0.45 |
|  |  | Student | 2.2% | 58 | 25.4% | 53 | 91.4<br>% | 5 | 8.6% | 0.57 | 0.20 –<br>1.58 | 0.28 |
|  |  | Housewife | 3.9% | 69 | 30.3% | 60 | 87.0<br>% | 9 | 13.0<br>% | 1.04 | 0.44 –<br>2.42 | 0.92 |
|  |  | Businessman | 0 | 3 | 1.3% | 3 | 100<br>% | 0 | 0 | NA |  |  |

In contrast, no statistically significant association was found between occupation and mortality. These findings suggest that lower educational attainment may be linked to poorer outcomes, potentially due to delayed care-seeking behavior, limited understanding of toxic exposure, or difficulty in recognizing early warning signs.

When the mortality in relation to OP and non-OP was compared, it was found that GCS<9, pupillary size especially constricted pupil was found to have predicted value of fatality (Table 7)

**Table 7:**
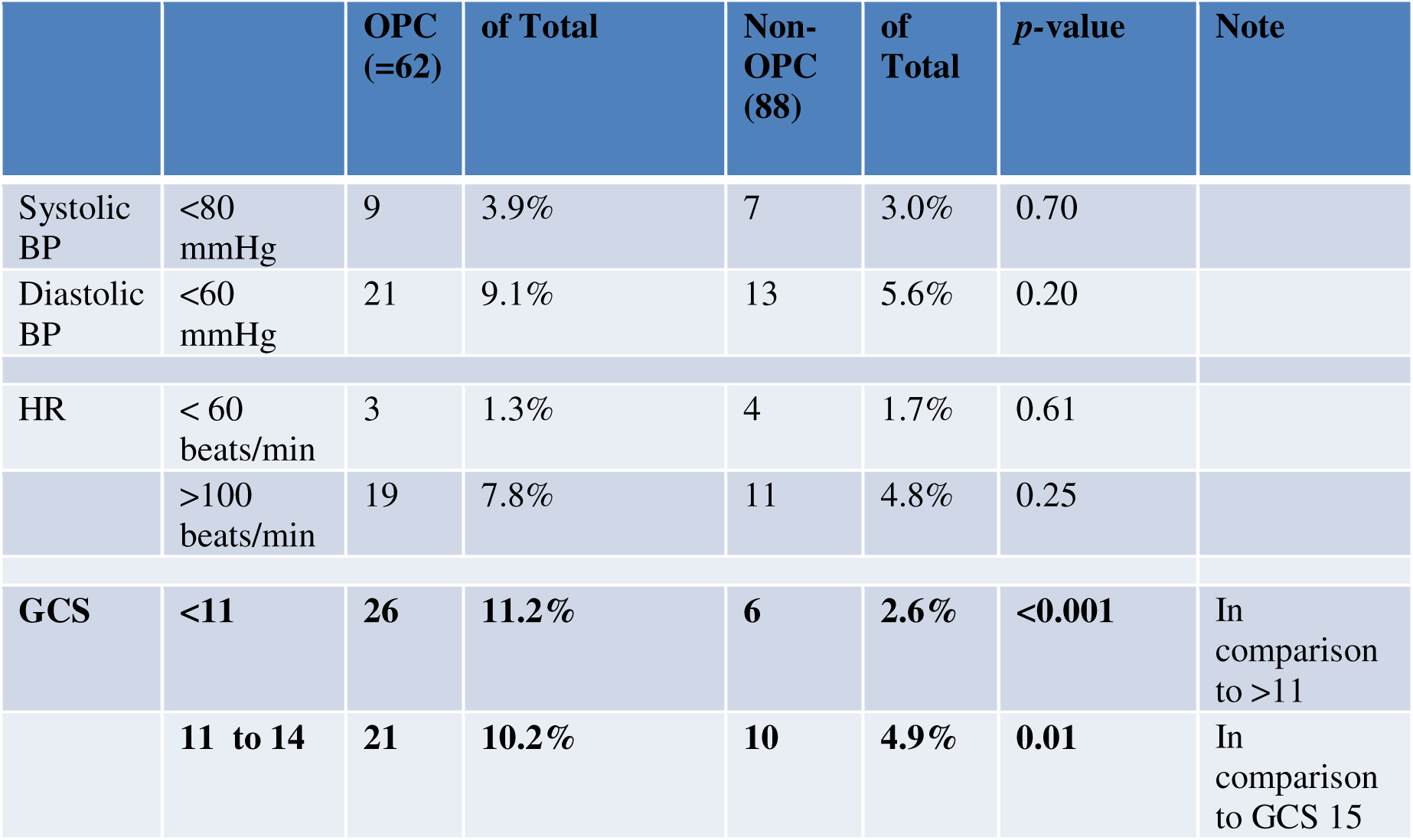
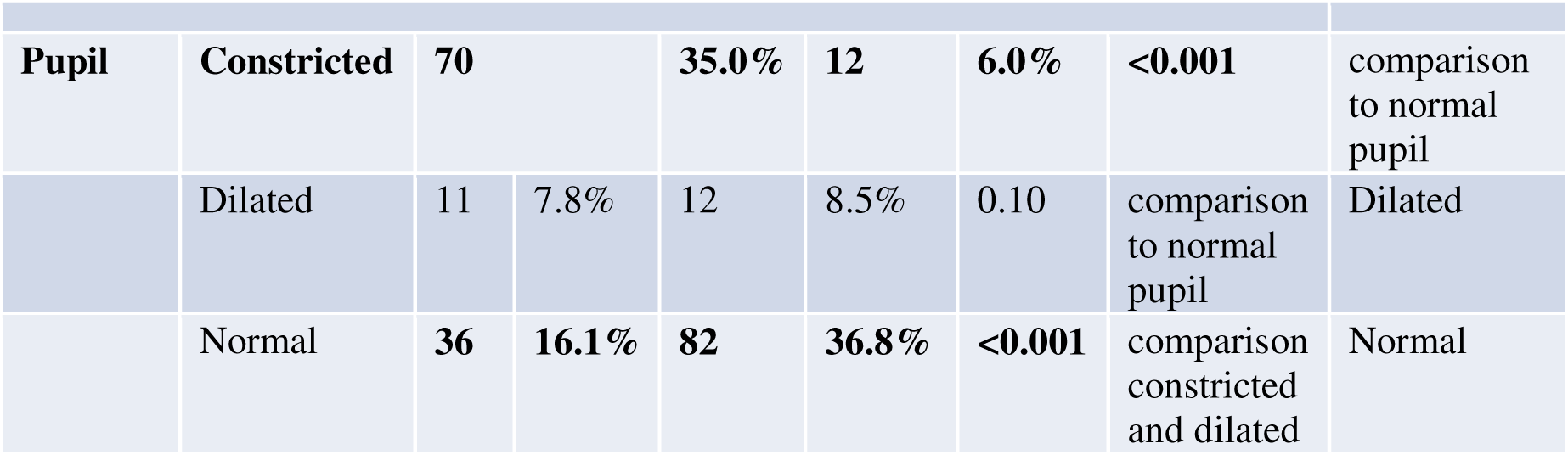
Comparison of vital signs and other features in between OP and non-OP poisoning.

|  |  | OPC<br>(=62) | of Total | Non-OPC<br>(88) | of<br>Total | p-value | Note |
| --- | --- | --- | --- | --- | --- | --- | --- |
| Systolic<br>BP | <80<br>mmHg | 9 | 3.9% | 7 | 3.0% | 0.70 |  |
| Diastolic<br>BP | <60<br>mmHg | 21 | 9.1% | 13 | 5.6% | 0.20 |  |
| HR | < 60<br>beats/min | 3 | 1.3% | 4 | 1.7% | 0.61 |  |
|  | >100<br>beats/min | 19 | 7.8% | 11 | 4.8% | 0.25 |  |
| **GCS** | **<11** | **26** | **11.2%** | **6** | **2.6%** | **<0.001** | In<br>comparison<br>to >11 |
|  | **11 to 14** | **21** | **10.2%** | **10** | **4.9%** | **0.01** | In<br>comparison<br>to GCS 15 |

|  |  |  |  |  |  |  |  |  |
| --- | --- | --- | --- | --- | --- | --- | --- | --- |
| **Pupil** | **Constricted** | **70** |  | **35.0%** | **12** | **6.0%** | **<0.001** | comparison to normal pupil |
|  | Dilated | 11 | 7.8% | 12 | 8.5% | 0.10 | comparison to normal pupil | Dilated |
|  | Normal | **36** | **16.1%** | **82** | **36.8%** | **<0.001** | comparison constricted and dilated | Normal |

Variable factors in causality and motive were observed in surviving and death cases of pesticide poisoning. Among the suicidal attempt cases, failing to pass exam (OR 1.47) and economic loss (OR 2.45) was found significance in committing deliberate self-harm. (Table 8)

**Table 8:**
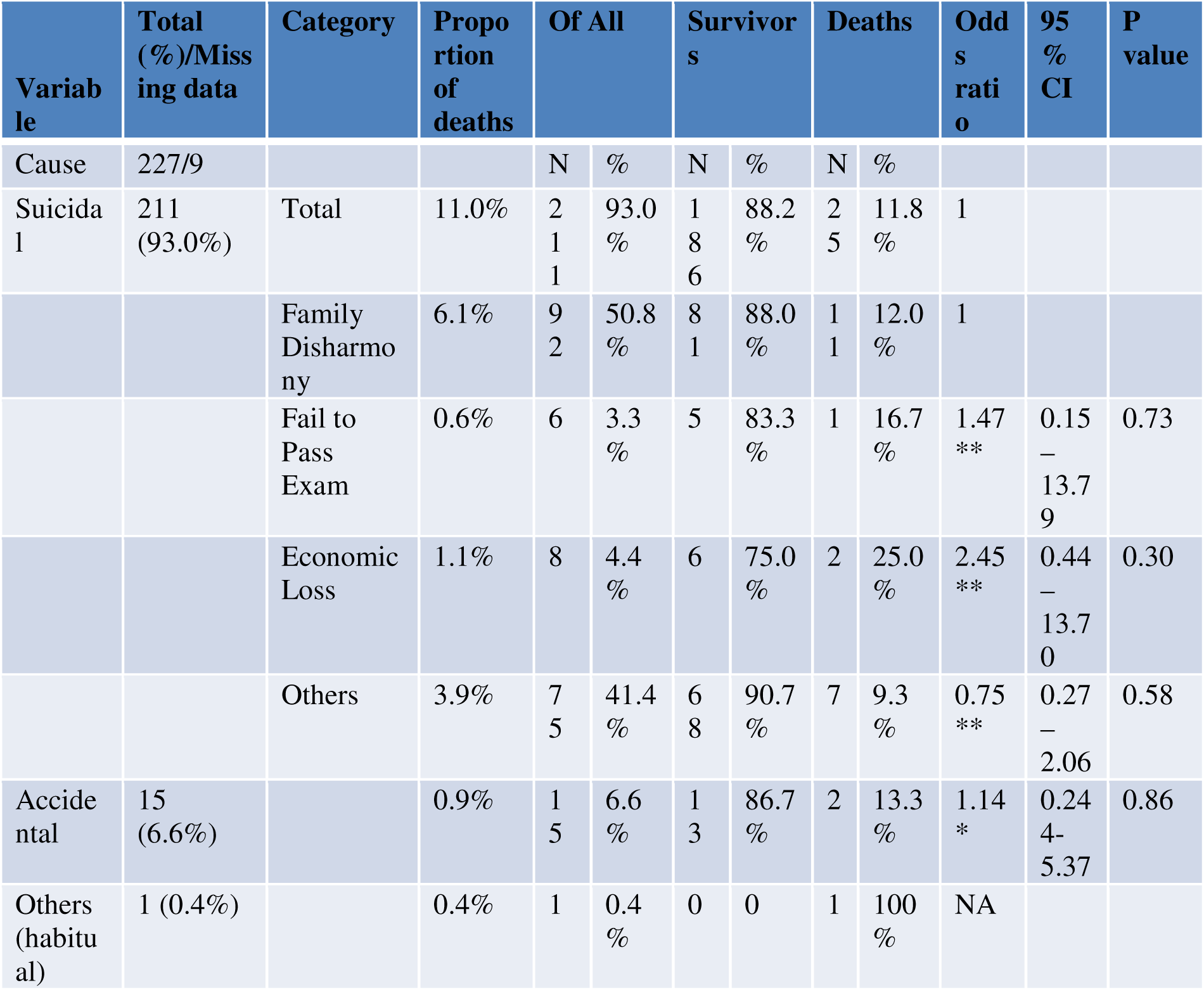
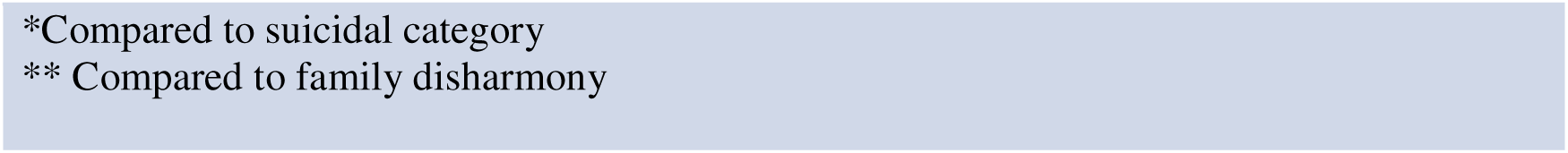
Odds Ratio (OR) of cause and motives of poisoning in causing death of patients.

| **Variable** | **Total (%) / Missing data** | **Category** | **Proportion of deaths** | **Of All** |  | **Survivors** |  | **Deaths** |  | **Odds ratio** | **95 % CI** | **P value** |
| --- | --- | --- | --- | --- | --- | --- | --- | --- | --- | --- | --- | --- |
| Cause | 227/9 |  |  | N | % | N | % | N | % |  |  |  |
| Suicidal | 211 (93.0%) | Total | 11.0% | 211 | 93.0 % | 186 | 88.2 % | 25 | 11.8 % | 1 |  |  |
|  |  | Family Disharmony | 6.1% | 92 | 50.8 % | 81 | 88.0 % | 11 | 12.0 % | 1 |  |  |
|  |  | Fail to Pass Exam | 0.6% | 6 | 3.3 % | 5 | 83.3 % | 1 | 16.7 % | 1.47 ** | 0.15 – 13.79 | 0.73 |
|  |  | Economic Loss | 1.1% | 8 | 4.4 % | 6 | 75.0 % | 2 | 25.0 % | 2.45 ** | 0.44 – 13.70 | 0.30 |
|  |  | Others | 3.9% | 75 | 41.4 % | 68 | 90.7 % | 7 | 9.3 % | 0.75 ** | 0.27 – 2.06 | 0.58 |
| Accidental | 15 (6.6%) |  | 0.9% | 15 | 6.6 % | 13 | 86.7 % | 2 | 13.3 % | 1.14 * | 0.24 – 5.37 | 0.86 |
| Others (habitual) | 1 (0.4%) |  | 0.4% | 1 | 0.4 % | 0 | 0 | 1 | 100 % | NA |  |  |

\*Compared to suicidal category \*\* Compared to family disharmony

OP was found to have the highest mortality (8.8%) among all cases of registered cases of pesticide. The herbicide followed with 3.1% cases although among the single cumulating cases of herbicide showed 33% death cases (OR 2.47). (Table 9)

**Table 9:** Proportion and mortality of type of poisoning (with logistic regression for mortality)

| Variable | Of All |  | Survivors |  | Deaths |  | Mortality | Odds ratio | 95% CI | P value |
| --- | --- | --- | --- | --- | --- | --- | --- | --- | --- | --- |
| Total/Missing data – 228/8 | N | % | N | % | N | % |  |  |  |  |
| OP | 119 | 52.2% | 99 | 83.2% | 20 | 16.8% | 8.8% | 1 |  |  |
| Carbamate | 9 | 3.9% | 9 | 100.0% | 0 | 0 | 0 | NA |  |  |
| Stored grain pest | 16 | 7.0% | 14 | 87.5% | 2 | 12.5% | 0.9% | 0.70* | 0.14 – 3.35 | 0.66 |
| Other pesticide/Miticide | 48 | 21.1% | 47 | 97.9% | 1 | 2.1% | 0.4% | 0.15* | 0.01-0.80 | 0.03 |
| Herbicide | 21 | 9.2% | 14 | 66.7% | 7 | 33.3% | 3.1% | 2.47* | 0.88 – 6.91 | 0.84 |
| Fungicide | 1 | 0.4% | 1 | 100% | 0 | 0 | 0 | NA |  |  |
| Rodenticide | 6 | 2.6% | 6 | 100% | 0 | 0 | 0 | NA |  |  |
| Other | 8 | 3.5% | 8 | 100% | 0 | 0 | 0 | NA |  |  |

\* Compared to OPC

Among the organophosphate (OP) compounds identified in this study, chlorpyrifos was associated with the highest mortality rate, accounting for 9.1% of total deaths (Table 10). Other OP agents, including combination products containing chlorpyrifos and cypermethrin, as well as malathion and phenthoate, each accounted for a mortality rate of 1.8%. Interestingly, dimethoate, which is often cited in the literature as a high-risk compound, was linked to a relatively low mortality rate of 0.9% in this cohort. These findings suggest variability in clinical outcomes based on the specific OP agent ingested, possibly influenced by dose, formulation, or time to treatment.

**Table 10:**
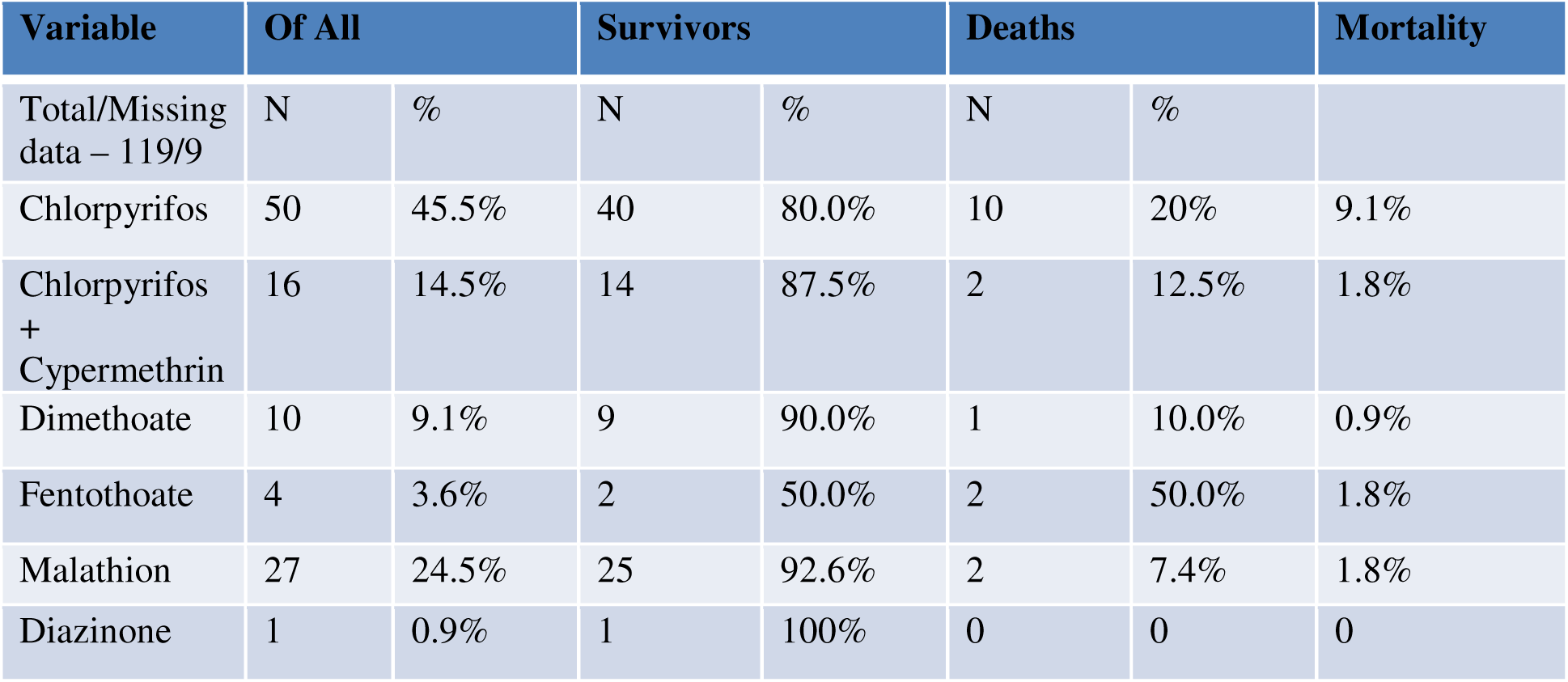
Proportion and mortality of type of OP compound.

Based on the confirmed specimens collected during the study, both organophosphate (OP) and non-organophosphate (non-OP) compounds were identified and catalogued according to their generic names, trade names, and active ingredients (Tables 11 and 12).

**Table 11.** : OPC and Carbamate identification list.

| Agphos | Chlorpyriphos | Organophosphate |
| --- | --- | --- |
| Basodin | Diazinon | Organophosphate |
| Chita | Chlorpyriphos | Organophosphate |
| Chlorban | Chlorpyrifos | organophosphate |
| Cithion | Malathion | Organophosphate |
| Classic | Chlorpyrifos | Organophosphate |
| Corolux | Quinalphos | Organophosphate |
| cyperfos | Chlorpyriphos + cypermethrin | Organophosphate + Pyrethroid |
| Diazinon | Diazinon | Organophosphate |
| Dimegro | Dimethoate | Organophosphate |
| Diphion | Malathion | Organophosphate |
| filnen | Diazinon | Organophosphate |
| Furadan | Carbofuran | Carbamate |
| Fyfanon | Malathion | Organophosphate |
| Glorious | Chlorpyrifos | Organophosphate |
| Hiltheon | Malathion | Organophosphate |
| Kap | Phenthoate | Organophosphate |
| Kinolux | Quinalphos | Organophosphate |
| Malathion | Malathion | Organophosphate |
| Marshall | Carbosulfan | Carbamate |
| Morter | Chlorpyrifos | Organophosphate |
| Nitro | Chlorpyrifos + cypermethrin | Organophosphate + Pyrethroid |
| Razhdon | Diazinon | Organophosphate |
| Ribbon | Chlorpyrifos | Organophosphate |
| Rocket | Prophenophos + cypermethrin | Organophosphate + Pyrethroid |
| Sabion | Diazinon | Organophosphate |
| Sangor | Dimethoate | Organophosphate |
| selection | Chlorpyrifos + cypermethrin | Organophosphate + Pyrethroid |
| semicron | Prophenophos + cypermethrin | Organophosphate + Pyrethroid |

**Table 12:**
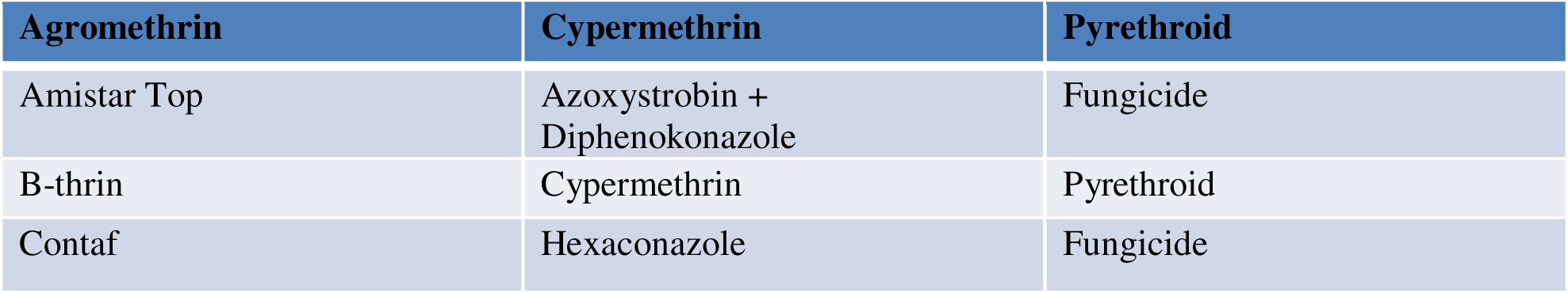
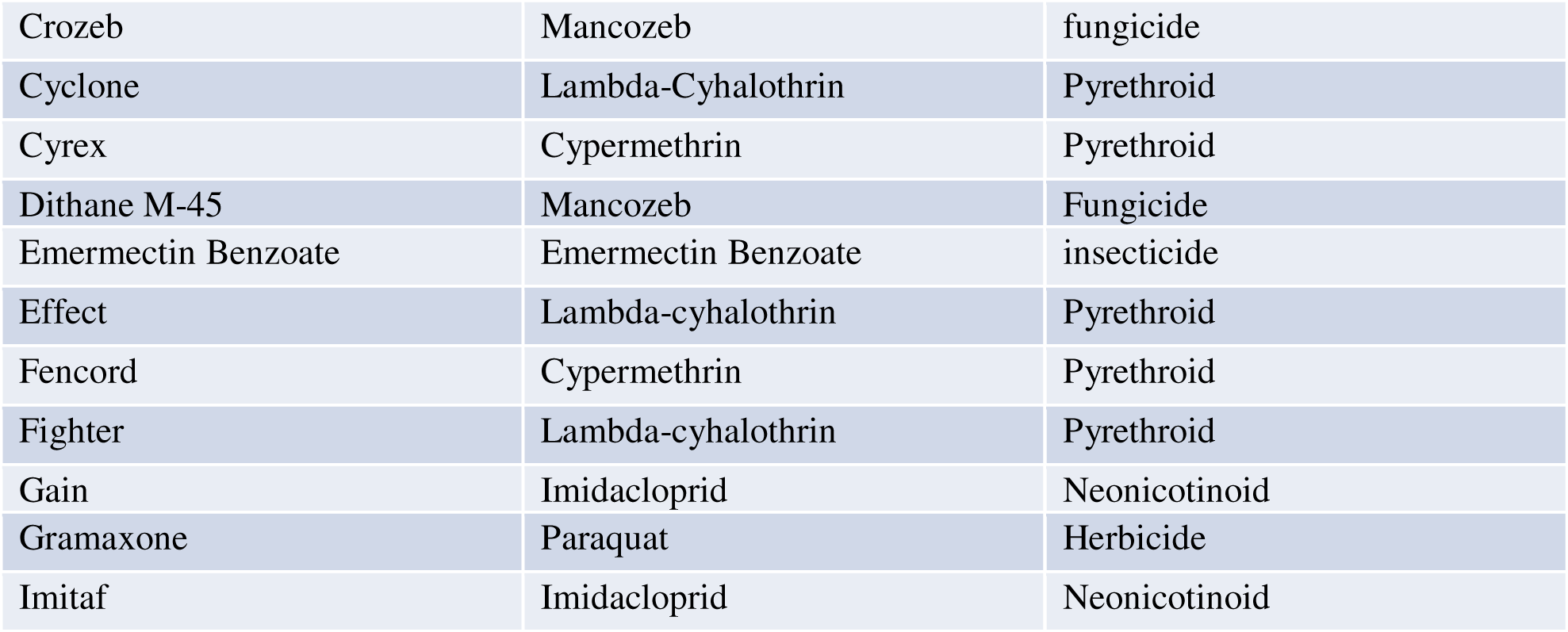
Non-organophosphate pesticide list.

These comprehensive reference tables offer valuable practical utility in clinical settings, particularly in resource-limited environments where toxicological analysis is unavailable. The information can assist clinicians, especially junior doctors, in making rapid, evidence-based treatment decisions when a pesticide container or label is presented.

The compilation underscores the importance of encouraging patients or attendants to bring physical samples or clear images to healthcare facilities, as this simple practice can significantly improve diagnostic accuracy and management outcomes. Visual tools, such as pictorial charts or clipboard references derived from these tables, may serve as effective bedside aids in emergency and internal medicine wards.

## Discussion

This cross-sectional study analyzed 236 cases of acute pesticide poisoning admitted to the Department of Medicine at Dhaka Medical College Hospital (DMCH), Bangladesh. Organophosphate (OP) compounds were the most frequently implicated agents in deliberate self-harm, which aligns with findings from previous studies conducted in similar regional contexts ^5,6,7,8,9^. Pyrethroids were identified as the second most common group. The case fatality rate was notably higher among patients exposed to OP compounds (10.9%) compared to those exposed to non-organophosphate substances. However, it is important to note that non-OP agents were responsible for approximately one-third of all deaths in this study cohort. These findings highlight an urgent need for broader, population-based research to evaluate the mortality burden from non-OP compounds such as stored grain pesticides and herbicides, demonstrating disproportionately high fatality rates in this series.

Although World Health Organization (WHO) Class I agents ^10^ less frequently encountered, cases involving their combination with OPs or pyrethroids were associated with severe toxicity and poor outcomes, while OPs remain the leading cause of mortality, specific non-OP agents, especially herbicides and grain storage pesticides, warrant special attention, as limited safer alternatives are currently available. Most poisoning cases in this study were managed in public hospitals. This may be due to the medicolegal implications of poisoning, which may deter private facilities from accepting such cases, even those requiring critical care. A similar treatment pattern has been observed in Sri Lanka, where public institutions manage most poisoning cases ^11^.

The incidence of poisoning was higher among males than females, consistent with national and international data ^4,9,12^. Young males aged 15–25 years represented the largest demographic group affected by deliberate self-poisoning. This finding supports the hypothesis that younger individuals may be more prone to impulsive behavior under acute psychosocial stress, as noted in prior studies ^3,4^. These results challenge the assumption that pesticide self-poisoning is primarily premeditated among older adults. A substantial proportion of patients (92%) were classified as suicidal in intent. Of these, 60% had completed only primary education, while 40% were illiterate. There was no significant variation in poisoning incidence based on religious affiliation. Most patients were married and from rural areas, reflecting known vulnerability patterns reported across South Asia, including in India and Sri Lanka ^2,6,11^.

Nausea and vomiting were the most reported symptoms. Significantly, the low Glasgow Coma Scale (GCS) score at presentation was strongly associated with poor outcomes. While the use of toxidrome-based clinical diagnosis was relatively effective for identifying OP poisoning, it was less reliable for non-OP compounds ^13^. This finding underscores the necessity for individualized assessment and, where possible, laboratory confirmation to improve diagnostic accuracy in pesticide-related poisoning.

A key strength of this study was the practical approach of prompting patients and their attendants to bring samples of the ingested substances either physically or via mobile-based images. The nearly equal distribution between OP and non-OP agents further reinforces the clinical utility of this method. Proper sample identification facilitates appropriate and targeted management and helps prevent unnecessary or excessive antidote use, such as atropine, thereby reducing treatment-related complications. Moreover, improved identification may contribute to earlier discharge and reduced healthcare costs.

### Study limitation

This study was conducted at a tertiary care hospital, which may not accurately represent Bangladesh’s broader epidemiology of pesticide poisoning. Since most poisoning cases are initially present in primary and secondary healthcare facilities, future research should focus on these settings to capture the full range of poisoning incidents. Moreover, the lack of laboratory-based toxicological confirmation limits the ability to validate clinical diagnoses. The findings underscore the need for comprehensive nationwide surveys to determine the true incidence, prevalence, morbidity, and mortality associated with pesticide poisoning in Bangladesh.

### Recommendation

A national poisoning registry is recommended to improve the management and surveillance of poisoning cases, beginning with major referral hospitals such as DMCH. Furthermore, a centralized Poison Information Center should be developed, equipped with a 24/7 call service to provide real-time guidance to healthcare providers managing toxicological emergencies. Public health campaigns are also guaranteed to promote early sample collection and timely hospital presentation.

## Conclusion

This study found that cases of pesticide poisoning were nearly evenly split between organophosphate (OP) and non-organophosphate (non-OP) types. It is essential to differentiate between these two categories early on, as their clinical management differs significantly. Unfortunately, many non-OP poisoning cases were initially treated using OP protocols, which may have resulted in inappropriate or excessive treatment and potential side effects. To help clinicians identify the specific agent involved and tailor the appropriate treatment, it is beneficial to encourage patients or their attendants to bring physical samples or photographs of the substances they ingested. Early identification supports targeted clinical care and may help reduce complications, resource usage, and overall treatment costs.

## Data Availability

All data produced in the present work are contained in the manuscript

